# Sanitation Practices and Child Health Outcomes in Gulu District: The Moderating Effect of Climate, Age, and Water Access

**DOI:** 10.64898/2026.05.29.26354417

**Authors:** Yoweri Idiba, Norman David Nsereko, Alex Barakagira

## Abstract

**Background:** The sanitation crisis poses a significant public health risk, leading to diseases like diarrhoea, cholera, and typhoid, which impede children’s health and development in developing countries like Uganda. Improving sanitation infrastructure is crucial for safeguarding child health and future generations. However, the link between sanitation and children’s health is complex, influenced by various factors. This investigation in Gulu scrutinises the correlation between sanitation practices and child well-being, considering moderating factors such as age, climate, and consistent water accessibility.

**Methods:** The study used a convergent parallel design with equal priority. The Social Ecological Model, Social Learning Theory, and Diffusion of Innovations Model guided it. Researchers collected data from 10 health facilities and 317 households, using purposive and simple random sampling. They used sampling proportions proportional to village size within strata. The researcher analysed quantitative data using SPSS with factor analysis, structural equation modelling, and multivariate analysis. To analyse qualitative data, they used DQA Minor Lite software, which facilitated thematic analysis.

**Results:** The finding shows 56.8% of households had low socio-economic status. Sanitation was poor; 24.9% household had improved latrines, 20.5% had handwashing facilities with soap, and 68.1% used basic anal cleansing. For nutrition, 38.5% of children were malnourished by MUAC; by Z-scores, 28.7% were stunted, 16.4% underweight, 13.6% wasted. Diarrhoea affected 62% of children. Climate worsened sanitation: 48.3% had latrines collapse from floods, and 63.4% of waterborne diseases occurred in both dry and wet seasons. Moderation analysis on childhood diarrhoea shows that sociocultural factors (β = -0.20, p < 0.001), sanitation (β = -0.15, p < 0.001), and health system response (β = -0.18, p < 0.001) reduced diarrhoea. Climate change increased risk (β = 0.15, p < 0.001) and moderated sanitation effects (β = 0.01, p < 0.05). Models explained 10–14% variance. Age and water access had no moderating effect. While childhood malnutrition shows that sociocultural factors (β = -0.43, p < 0.001) and health system response (β = -0.13, p < 0.001) reduced malnutrition. Sanitation had no effect (β = 0.01, p > 0.05). Age increased malnutrition risk (β = 0.28, p < 0.01) and moderated sociocultural effects (β = 0.16, p < 0.001), but not sanitation. The model explained 21% variance, R² = 0.21, p < 0.001.

**Conclusion:** Sociocultural improvements and health system responses lower both diarrhoea and malnutrition. Climate worsens diarrhoea and alters sanitation’s impact. Age worsens malnutrition and changes sociocultural effects. These findings are valuable for policymakers, healthcare professionals, and researchers

## I. INTRODUCTION

Access to safe sanitation is a major global health challenge, and its availability varies across countries (WHO & UNICEF, 2023). While Sub-Saharan Africa has made progress in reducing open defecation, issues remain. Climate plays a role in shaping sanitation practices and their health outcomes. The connection between sanitation and child health is complex. Factors such as age, climate, and water access act as moderators in this relationship. Health system operations and established cultural customs influence these elements (Biran et al., 2022). The study examines how climate, age, and access to safe water interact with sanitation practices to impact child health - diarrhoea and malnutrition, considering the influence of cultural norms and local health infrastructure. The study examined what influences caregivers’ views on sanitation and found that community-led efforts lead to improvements. It also examines the healthcare system’s provision of quality, accessible services. It also considers the risks of improper waste disposal, especially contaminating water sources (Bauza & Guest, 2017).

Open defecation and unsafe waste disposal are common poor sanitation practices in low-income countries. These practices elevate the risk of diarrhoeal diseases, as noted by Contreras et al. (2022) and have a negative impact on child health. Studies show that resource-limited areas require new health strategies to address these issues. Understanding the social and economic reasons behind sanitation access, especially for children under five, is key (Islam et al., 2020). Ultimately, addressing these disparities is paramount to develop effective public health strategies.

Focusing on East Africa, Uganda inclusive, highlights the severe sanitation and health challenges children face (Delahoy et al., 2018). Vulnerable populations suffer from diseases linked to poor sanitation, worsened by systemic failures. In Gulu District, 28% of households still practice open defecation, and problems with diaper waste and a lack of clean water put children’s health at risk. Diarrhoea, malaria, and respiratory illnesses are the leading causes of death, according to the UBOS-UDHS (2022) and the 2023 Gulu District Health Officer report.

Understanding the connection between sanitation and health requires focusing on how different age groups are affected. Pickbourn and Ndikumana (2022) highlighted that infants and young children are vulnerable to sanitation-related illnesses because of their developing immune systems and behaviours such as hand-to-mouth contact. Urgent interventions are needed to address age-specific risks and promote early childhood hygiene, emphasizing age-related sanitation vulnerabilities.

Poor hygiene and sanitation make toddlers (1-3 years old) susceptible to diarrhoea and malnutrition, as noted by WHO and UNICEF (2023). Their natural curiosity and love of exploration increase the risk of swallowing faeces or contaminated water (Bureau for Children, 2023). Children aged 4-12 are vulnerable to waterborne illnesses and parasites from unsafe water/conditions (WHO, 2018); Bureau for Children, 2023). Sanitation programs adapt to different age groups, recognising the specific risks children encounter as they grow.

Climate factors influence sanitation practices and their resulting health outcomes. As noted by Van Meyel et al.(2022), climate variations in temperature and rainfall influence sanitation and hygiene. When areas get hit by extreme weather like floods, their sanitation systems break, which affects children’s health. The link highlights the critical need for sanitation that handles changing environmental conditions. Detailed research lacks on severe weather’s impact on hygiene and health. Studies by Dhimal et al.(2022) and Okesanya et al.(2024) stress prizing adaptive strategies suited to local environmental contexts. A gap remains in research that explores these dynamics.

Safe water is vital. It affects sanitation and child health. Shrestha et al.(2020) suggest combining data on water access and sanitation. This helps understand their health effects. Rural children face disease risks. The issue arises from scarce potable water. Future research will examine these connections. Despite recent progress, major gaps remain in current research. Pickbourn et al.(2022) highlight infancy and early childhood as crucial periods for sanitation-related health risks. However, there is limited investigation into age-specific vulnerabilities across different developmental stages. Therefore, future longitudinal studies must investigate how sanitation interventions can be best adapted to the specific needs of children at various developmental stages.

The implications of climate change for sanitation infrastructure and practices warrant further examination. Current research does not explain how climate change affects sanitation and health. Current research highlights the pressing need for context-specific, adaptive strategies to enhance climate resilience. The connection between safe water access and sanitation practices represents a major research gap. Shrestha et al.(2020) noted data integration challenges in water and sanitation, stressing the need for studies on water access, sanitation, and child health.

Future research will explore key areas. It will discuss aging risks. Researchers will examine climate change’s effects on sanitation and children’s health. Researchers will cover the benefits of combining safe water and sanitation for public health. A comprehensive plan is necessary. This plan will fight sanitation’s health dangers. It will also improve children’s health worldwide.

## II. MATERIALS AND METHODS

### Study Design

The study employed a convergent parallel design, assessing data at the same time and valuing both viewpoints. Using quantitative and qualitative approaches, the study explored the influence of climate, age, and water on children’s sanitation habits and health outcomes (Creswell & Clark, 2017). Using a pragmatic mixed-methods research framework, the investigation examined the topic’s complexity and showed the benefits of integrating quantitative and qualitative methods (Morgan, 2007; This research significantly enhances understanding of how age, climate, and water access influence the relationship between sanitation practices shaped by diverse sociocultural and systemic factors and child health outcomes. These outcomes offer major insights for creating focused public health initiatives and measures that align with local demands and settings.

### The Study Context

A study was conducted in Awach and Owoo sub-counties, Gulu District, Uganda, to examine how sanitation affects childhood diarrhoea and malnutrition. These sub-counties were selected for their distinct geographical and demographic features. Gulu District has a tropical climate, varied terrain, and a population of 135,373 (UBOS 2024), making it ideal for sanitation studies. Awach sub-county has 13,002 people (6,371 males, 6,631 females) in 2,167 households. Owoo sub-county has 13,506 people (6,618 males, 6,888 females) in 2,251 households, giving a total target population of 4,418 households (UBOS 2024). The accessible population included 3,399 household caregivers, healthcare providers, and community leaders. Participants had to be aged 18+, residents of the sub-counties, and able to give informed consent. The study used a multilevel approach combining quantitative and qualitative data, considering age, climate, and water access as key factors influencing sanitation practices, diarrhoea, and malnutrition in children.

### Recruitment and Participant Selection

The study employed a multistage sampling design that combined probability and nonprobability sampling. Fisher’s formula (n = 317) helped researchers calculate the quantitative sample size. For the qualitative part, used data saturation to determine the sample size (n = 43). In this context, n stands for the sample size, Z for the standard normal deviation at 1.96 for a 95% confidence level, P for the proportion of the target population with diarrhoea (which is 29.1%, based on a study by (Omona et al., 2020), q for 1–P, and d for the margin of error, set at 0.05 (5%) for households and 0.1 (10%) for villages. This formula’s selection used study-relevant reference points.

Maximum variation sampling ensured participant diversity. Classifying sub-counties considered rural or urban status, with a focus on those with low sanitation coverage. We selected villages and households using simple random sampling. The sampling proportions within each stratum were proportional to size. From each stratum, researchers purposively selected participants for the focus group discussions (FGDs). The group comprised men and women who were both caregivers for children in their households and members of the sanitation committee. Expert sampling allowed to select key informants for in-depth interviews. Researchers recruited 360 participants. This group had 317 people. They were household caregivers of children under five. It also included 33 community members. These members participated in four focus groups. Sanitation committee members were part of these groups. Researchers also included ten key informants. Various health professionals, including doctors, nurses, and educators, as well as health officials and sanitation managers from both community groups and the Ministry of Health, took part in key interviews.

### Data Collection and Ethical Consideration

Data collection was conducted from 21st October 2024 to 15th December 2024. Prior to data collection, written informed consent was obtained from all adult participants. For households with children under five, parental and guardian consent was secured, and minor assent was obtained where age-appropriate, in line with ethical guidelines for research involving children. Participants were briefed on the study purpose, procedures, risks, and benefits, and were assured of confidentiality and anonymity. They were informed of their right to withdraw at any time without penalty, and pseudonyms were used for qualitative data. The study adhered to the Declaration of Helsinki and received ethical approval from the Nkumba University Vetting Committee, Lacor Hospital Institutional Review Board, Uganda National Council for Science and Technology, and district officials.

Structured questionnaires designed by the researcher and WHO were used to collect quantitative data on demographics, sanitation, climate change, water access, child diarrhoea, and malnutrition. Trained research assistants surveyed 317 households with children under five and conducted sanitation observations. The Local Council supported sampling by providing household lists and village maps. Each household survey lasted for 60 minutes, based on pre-tests in similar settings.

For qualitative data, focus group discussions were held with caregivers, sanitation committee members, and community leaders in groups of 8–10, organized by health inspectors at sub-county facilities. U-shaped seating was used to encourage participation. In-depth interviews with healthcare professionals, sanitation officers, and community leaders were conducted by the lead author in Acholi with translator support. The FGDs and interviews were audio-recorded, transcribed verbatim, and translated into English. Discussions explored links between age, climate, water access, sanitation, child health, and caregiver views.

Data collection occurred in private settings with strict access controls and secure storage to protect sensitive information. To ensure consistency, different research assistants were assigned to each Subcounty, and separate FGDs were held with caregivers and sanitation committee members.

#### Validity and reliability

Quantitative data validity was established through face validity assessment. Confirmatory Factor Analysis showed good model fit, and Structural Equation Modelling revealed significant associations between sociocultural norms and childhood diarrhoea/malnutrition, moderated by socioeconomic status and sanitation access. Reliability was confirmed with Cronbach’s alpha of 0.85. Qualitative rigor was ensured through member checking, peer debriefing, prolonged engagement, data triangulation, and inter-rater/intra-rater reliability checks. Quantitative and qualitative data were merged during interpretation.

### Data Analysis Procedure

A sequential mixed-methods approach was used to analyse both quantitative and qualitative data, ensuring a comprehensive understanding of the research problem. Quantitative data were analysed using SPSS and Jamovi, while qualitative data were examined through deductive thematic analysis in QDA Miner Lite. Findings from both strands were later merged using abductive integration and triangulation to strengthen interpretation and test theory across contexts.

Quantitative analysis began with thorough data cleaning to address missing values, outliers, and entry errors. Exploratory Factor Analysis was first conducted to identify the underlying structure of the data, revealing a two-factor solution that explained 20.35% of the variance in cultural beliefs and educational knowledge. Confirmatory factor analysis was then applied to validate this model, demonstrating good fit and confirming both convergent and discriminant validity. With the measurement model established, descriptive statistics summarised key variables related to demographics, sanitation, water access, and child health outcomes. Bivariate logistic regression explored associations between sanitation facilities, diaper disposal practices, sociocultural factors, health system responses, and child diarrhoea and malnutrition. Variables significant at p < 0.05 were carried into multivariate logistic regression to adjust for confounders. Multicollinearity was checked using Variance Inflation Factors, with values kept below 10, and model fit was assessed with the Hosmer-Lemeshow test. Odds ratios with 95% confidence intervals were reported. Structural Equation Modelling tested hypothesised causal pathways among variables and validated the logistic regression results. Statistical significance was set at p < 0.05, and inductive reasoning guided interpreting quantitative findings.

For qualitative analysis; the focus group and interview transcripts were imported into QDA Miner Lite. A deductive coding framework was developed from the research objectives, literature review, and social-ecological model, with a detailed codebook defining each code and providing illustrative text examples. To ensure reliability, two research assistants coded a subset of transcripts, and discrepancies were resolved through discussion. The coded data were then examined to identify recurring patterns. Themes were developed based on the frequency and consistency of codes across transcripts and interpreted in relation to the study objectives and existing literature. This approach allowed sociocultural norms, sanitation practices, and caregiver perceptions to be re-evaluated within the social-ecological model.

In the final stage, quantitative and qualitative findings were integrated through abductive triangulation. This process enabled cross-validation of SEM pathways with caregiver narratives, highlighting areas of convergence and divergence between datasets. A comprehensive data management plan ensured integrity, reproducibility, and adherence to ethical and regulatory standards throughout analysis. By combining both methods, the study generated robust evidence to inform policy and practice on childhood diarrhoea and malnutrition.

## III RESULTS

### Socio-demographic characteristics of participants

Table 1 shows that 94% (298/317) of the caregivers were female. Among the children, 50.5% were male, and 50.5% were between 24 and 59 months old. 56.8% of households had low socioeconomic status. A proportion, 80.5% of households in the rural area of Owoo sub-county, had lower socioeconomic status than the proportion in the urban area of Awach sub-county (31.4%).

**Table 1:** Socio-demographic Characteristics and Sanitation Status.

| Variables | Categories | Awach Sub-county |  | Owoo Sub-county |  | Gulu District |  |
| --- | --- | --- | --- | --- | --- | --- | --- |
|  |  | Freq. | Percent | Freq | Percent | Freq | Percent |
|  |  | (n=153) | (%) | (n=164) | (%) | (n=317) | (%) |
| Caregiver gender | Male | 10 | 6.5 | 9 | 5.5 | 19 | 6.0 |
|  | Female | 143 | 93.5 | 155 | 94.5 | 298 | 94.0 |
| Child gender | Male | 79 | 51.6 | 81 | 49.4 | 160 | 50.5 |
|  | Female | 74 | 48.4 | 83 | 50.6 | 157 | 49.5 |
| Child age | Less 12 months | 39 | 25.5 | 38 | 23.2 | 77 | 24.3 |
|  | 13-23 months | 46 | 30.1 | 33 | 20.1 | 79 | 24.9 |
|  | 24-59 months | 68 | 44.4 | 93 | 56.7 | 161 | 50.8 |
| Socioeconomic status | Low | 48 | 31.4 | 132 | 80.5 | 180 | 56.8 |
|  | Medium | 103 | 67.3 | 31 | 18.9 | 143 | 45.1 |
|  | High | 2 | 1.3 | 1 | 0.6 | 3 | 0.9 |
| Mid-Upper Arm Circumference (MUAC) | Less 11.4cm | 18 | 11.8 | 6 | 3.7 | 24 | 7.6 |
|  | 11.5-12.5cm | 65 | 42.5 | 33 | 20.1 | 98 | 30.9 |
|  | Above 12.5cm | 70 | 45.8 | 125 | 76.2 | 195 | 61.5 |
| Weight for Age (WAZ) Z Scores | <-2SD | 41 | 26.8 | 11 | 6.7 | 52 | 16.4 |
|  | 2SD-2SD | 110 | 71.9 | 147 | 89.6 | 257 | 81.1 |
|  | 3SD | 4 | 2.6 | 17 | 10.4 | 21 | 6.6 |
| Height for Age (HAZ) Z scores | <-2SD | 44 | 28.8 | 47 | 28.7 | 91 | 28.7 |
|  | >-2SD to 3SD | 109 | 71.2 | 117 | 71.3 | 226 | 71.3 |
| Weight for Height (WHZ) Z scores | <-2SD | 34 | 22.2 | 9 | 5.5 | 43 | 13.6 |
|  | >-2SD to 3SD | 119 | 77.8 | 155 | 94.5 | 274 | 86.4 |
| Diarrhea 6months | Yes | 100 | 65.4 | 98 | 59.8 | 198 | 62.5 |
|  | No | 53 | 34.6 | 66 | 40.2 | 119 | 37.5 |
| Access to sanitation facilities | Yes | 115 | 75.2 | 123 | 75.0 | 238 | 75.1 |
|  | No | 38 | 24.8 | 41 | 25.0 | 79 | 24.9 |
| Hand washing Facilities | Yes | 39 | 25.5 | 26 | 15.9 | 65 | 20.5 |
|  | No | 114 | 74.5 | 138 | 84.1 | 252 | 79.5 |
| child excreta facilities | Yes | 97 | 63.4 | 66 | 40.2 | 163 | 51.4 |
|  | No | 56 | 36.6 | 98 | 59.8 | 154 | 48.6 |

The nutritional status of children showed that 38.5% were malnourished, showed by a Mid-Upper Arm Circumference (MUAC) of less than 12.5 cm. 7.6% were classified as severely underweight, with a MUAC below 11.5 cm. In the urban area of Awach, a higher percentage of undernourished children was reported at 54.3%, compared to 23.8% in Owoo sub-county. Using the World Health Organisation (WHO) standard S-scores, 28.7% of children stunted (HAS <-2 SD), 16.4% were underweight (WAS <-2 SD), and 13.6% wasted (WHS <-2 SD).

Diarrhoea affected 62.7% (199/317) of households across the sub-counties. This revealed that 24.9% (79/317) of households lacked access to sanitation facilities, and only 20.5% (65/317) practiced handwashing with soap. However, sharing latrines was common among these households. Half, 48.6%, of households lacked child excreta facilities, and 68.1% engaged in unimproved anal cleansing. Child toilets were available in 20.8% of homes with a slab, 32.2% without, 6.3% used potties/diapers, and the rest had open defecation.

Findings on soiled diaper disposal showed that 64.1% of households disposed of diapers in open spaces. Households achieved this by washing and reusing cloth diapers; however, 3.2% of households disposed of disposable diapers. 7.3% of households crudely wrapped and dumped diapers, whereas only 36% used pit latrines for disposal. Hygiene education was provided in 68.8% of households; potty or toilet training was observed in 40.1%; and guidelines on diaper use and disposal were shared in 43.5%, with no significant differences across sub-counties.

### Descriptive statistics on moderating factors: climate change, age, and access to water

According to Table 2, the research findings provide important insights into sanitation practices, child health, and the effects of climate change. It’s noteworthy that 60.6% (192/317) of participants stated sanitation habits decrease as children age, compared to 32.2% (102/317) who found they increase. This raises concerns about the potential neglect of essential hygiene as children age. Specific sanitation needs for children under five were identified: 47.0% (149/317) of respondents emphasized the need for separate sanitation facilities, and 38.2% (121/317) advocated for routine faecal disposal. This underscores an urgent need for age-appropriate sanitation solutions to promote the health and well-being of young children.

**Table 2:** Shows descriptive statistics on moderating factors.

| Variables | Categorical responses | Frequencies<br>(n=317) | Percentages<br>(%) |
| --- | --- | --- | --- |
| Changes in Sanitation Practices as children grow | Become more frequent | 102 | 32.2 |
|  | Become less frequent | 192 | 60.6 |
|  | Remain the same | 7.2 | 5.0 |
| Specific Sanitation Needs for Children Under 5 | More frequent disposal of faeces | 121 | 38.2 |
|  | Separate sanitation facilities | 149 | 47.0 |
|  | Increased access to safe water | 38 | 12.0 |
|  | Others | 9 | 2.8 |
|  | Diarrheal diseases | 199 | 62.8 |
| Age-specific health risks associated with poor sanitation | Intestinal worms(STH) | 46 | 14.5 |
|  | Respiratory infections | 29 | 9.1 |
|  | Water contamination | 22 | 6.9 |
|  | Skin diseases | 21 | 6.6 |
| Impact of Climate Change on Sanitation | Collapsed latrine due to flooding | 153 | 48.3 |
|  | Contaminated water | 93 | 29.3 |
|  | Outbreaks like dysentery, cholera | 33 | 10.4 |
|  | Others | 38 | 12.0 |
| Perceived Impact of Climate Change on Waterborne Diseases | Not impactful | 71 | 22.3 |
|  | Neutral | 45 | 14.2 |
|  | Significantly Impact | 201 | 63.4 |
| Climate-Related Issues Experienced by Children | Yes | 125 | 39.4 |
|  | No | 192 | 60.6 |
| Water Availability and Quality due to Climate Change | Yes | 154 | 48.6 |
|  | No | 163 | 51.4 |
| Households located near a water source | Yes | 204 | 64.4 |
|  | No | 113 | 35.6 |
| Water source types | Borehole | 220 | 69.4 |
|  | Springwell | 19 | 6.0 |
|  | Topwater | 22 | 6.9 |
|  | Shallow wells | 31 | 9.8 |
|  | Other streams | 25 | 7.9 |
| Access to clean water | Yes | 262 | 82.6 |
|  | No | 55 | 17.4 |
| Child malnutrition due to climate change | Yes | 88 | 27.8 |
|  | No | 229 | 72.2 |
| Child waterborne illness (diarrhoea) in the past year | Yes | 236 | 74.4 |
|  | No | 81 | 25.6 |
| Increases in vector-borne diseases like malaria due to climate change | Yes | 277 | 87.4 |
|  | No | 40 | 12.6 |

The participants in our F.G.Ds painted a vivid picture of the devastating impact of poor sanitation on their lives. “*Diarrheal diseases are a constant threat to our children’s health,” said one participant*. Indeed, the data revealed that a staggering 62.8% of respondents (199/317) identified diarrheal diseases as the leading health risk associated with poor sanitation. This alarming statistic underscores the urgent need for public health efforts to prioritise sanitation infrastructure and hygiene practices.

However, it extends beyond sanitation; climate change also plays an important role in exacerbating these issues. As one participant noted, *“Our latrines are collapsing because of flooding, and gets complicated every year*. The quantitative data aligns with this, where 48.3% of participants (153/317) reported that household latrines had collapsed because of flooding, and 63.4% (201/317) believes that climate change influences the spread of waterborne diseases.

The impact on children is of particular concern; a notable 74.4% of respondents (236/317) reported that their children had experienced waterborne illnesses in the past year, underscoring a critical link among sanitation, climate change, and child health. *As one participant lamented, “It is heartbreaking to see our children suffer like this. We need help to improve our sanitation and protect their health.”*

The study underscores the extent of climate-related issues impacting children, with 39.4% (125/317) experiencing heat stress. The results show that although 82.6%(262/317) of households have access to clean water, half reported changes in water availability or quality caused by climate change, 48.6% (154/317) of respondents. This highlights the need for strategies to improve water quality management and sanitation practices amid environmental changes.

The key informants in our study highlight the complex interconnections among sanitation practices, health risks, and climate change, emphasizing that these issues are linked. “*You cannot talk about sanitation without talking about climate change,” said one key informant. “It is a vicious cycle: poor sanitation leads to health risks, which are worsened by climate change, and then it comes back to affect the children.”*

Qualitative findings suggest that addressing these issues requires a comprehensive approach that extends beyond the provision of sanitation infrastructure alone. “*We need to engage with the communities, educate them about the importance of proper sanitation and hygiene, and empower them to take ownership of their health,” said another key informant*.

By working together, stakeholders reduce the risks linked to poor health practices and foster health futures for children. “*It is not just about building latrines; it is about changing behaviors and creating an environment that supports good health,” said a key informant*. Our study emphasizes the need for a holistic approach to tackling sanitation, climate change, and child health, and with community involvement and education, can make a real difference.

Figure 1 illustrates various ways communities adapt to address the challenges of climate change and sanitation. Among the 317 respondents, the most common adaptation was digging water channels and trenches, reported by 73 individuals (23.0%), reflecting proactive efforts to control water flow and prevent flooding. 53/317 (16.7%) respondents reported reconstructing latrines, while 39/317 (12.3%) shared latrines, highlighting communal strategies and potential access issues.

**Figure 1:**
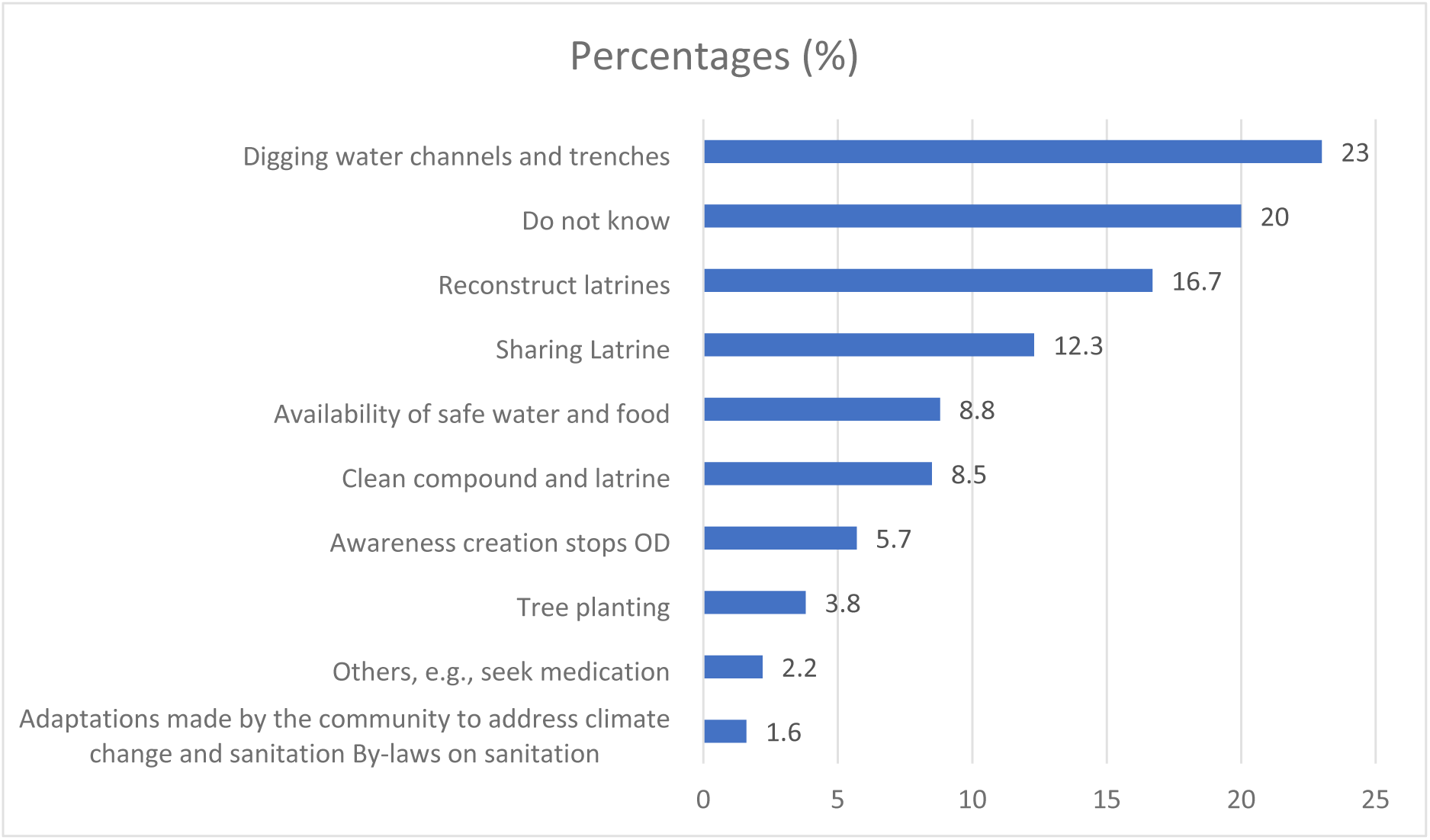
Climate change and sanitation adaptations

Awareness of the need to stop open defecation was reported by 18/317 (5.7%) respondents, showing recognition of the importance of education in improving sanitation practices. Other actions included cleaning compounds and latrines (27 respondents, 8.5%) and ensuring safe water and food were available (28 respondents, 8.8%). However, 55/317 respondents (17.3%) were unsure about which actions to take, underscoring the urgent need for improved education and resources.

Overall, although the community shows involvement in addressing sanitation and climate change, the results reveal gaps in knowledge and action that must be addressed to improve health outcomes. Targeted educational campaigns and support are crucial for empowering communities to adopt sustainable practices that improve sanitation and climate resilience.

### Data reduction through EFA, CFA, and SEM

Before conducting the moderation analysis, this study used exploratory factor analysis (EFA) and confirmatory factor analysis (CFA) to identify the underlying constructs related to sanitation and child health outcomes. EFA is a statistical technique used to detect patterns and relationships among variables, whereas CFA assesses the relationships between latent constructs and their indicators. The research questions and objectives guided the selection of variables for this study. The variables were selected based on their relevance to sanitation and child health outcomes, as well as their capacity to explain the relationships among these constructs.

#### Exploratory Factor Analysis (EFA)

The results of the Exploratory Factor Analysis (EFA) identify two main factors that enhance our understanding of the dataset’s underlying constructs. The factor loadings illustrate the relationship between each variable and the identified factors. Factor 1 is characterized by high loadings on water access and sanitation practices, while Factor 2 is defined by significant loadings on variables related to climate change and child health outcomes.

Table 3 shows that within Factor 1, variables such as W104 (loading = 0.698), W101 (loading = 0.550), and W108 (loading = 0.555) have strong positive loadings, showing their major contributions to this factor. For Factor 2, CM95 (loading = 0.690) and CM100 (loading = 0.679) also display high loadings. The factor statistics show that Factor 1 has a sum of squared loadings (SS Loadings) of 1.38, representing 15.3% of the variance. In contrast, Factor 2 has an SS Loading of 1.32, accounting for 14.7% of the variance. Combined, these factors account for 30% of the total variance, a substantial proportion consistent with standards for exploratory factor analysis.

**Table 3:** Factor load for moderating effects on sanitation and health outcomes.

| Item code | Code description | Factor loadings | Uniqueness | MO MSA |
| --- | --- | --- | --- | --- |
| CM95 | Climate change impacts on the spread of waterborne diseases | 0.545 | 0.690 | 0.661 |
| CM100 | Children experience climate-related health issues, -heat stress | 0.561 | 0.679 | 0.614 |
| W104 | Use cooling measures to protect the child from heat stress | 0.698 | 0.498 | 0.641 |
| W101 | Households located near a water source | 0.550 | 0.694 | 0.713 |
| W109 | Type of water source | 0.487 | 0.747 | 0.611 |
| W108 | Access to clean water | 0.555 | 0.674 | 0.667 |

Note. The 'principal axis factoring' extraction method was combined with a 'simplex' rotation, with CM representing climate and W representing water as the moderators.

Multiple indices support the appropriateness of the factor structure for assessing model fit. The RMSEA (Root Mean Square Error of Approximation) was 0.0767, which falls within the acceptable range for a good model fit; values below 0.08 are adequate. The TLI (Tucker-Lewis Index) score of 0.808 shows a reasonable fit, although values above 0.90 are preferred. The chi-squared statistic (χ² = 54.5, df = 19, p < .001) shows a significant difference between the observed and estimated covariance matrices, which is common in exploratory analyses that reflect complexity and estimated parameters. Assumption checks, including Bartlett’s Test of Sphericity, further support the validity of the factor analysis. The chi-squared value of 388 (p < 0.001) signifies a significant difference between the correlation matrix and the identity matrix, confirming the appropriateness of conducting factor analysis. The KMO measure of sampling adequacy yielded an acceptable overall value of 0.657, confirming the sample’s suitability for factor analysis.

In summary, exploratory factor analysis identifies two main factors that account for 30% of the variance in the dataset. The model shows a good fit, supported by suitable goodness-of-fit indices and validation of assumptions. The substantial factor loadings suggest variables reflect the underlying constructs, while the uniqueness values show variables are separate from the identified factors. These findings offer a solid basis for future confirmatory analyzes to verify the structure of the identified factors.

#### Confirmatory Factor Analysis

The Confirmatory Factor Analysis (CFA) results reveal a complex landscape, including factor loadings and overall model fit. The factor loadings show indicators are associated with their respective latent constructs, whereas others exhibit weak or negative associations. The covariance analysis reveals meaningful relationships among the factors, highlighting complex interactions among them. For example, the Climate Change factor is characterized by a significant loading on indicator CM 100 (0.451, p = 0.002), showing a moderate positive link. In contrast, indicators within the water factor have negative loadings for W102 (-0.876, p < .001) and W103 (-1.639, p < .001), which affect the water construct. The covariance analysis also reveals significant relationships among the constructs. Specifically, the positive covariance with the Climate Change factor (0.4715, p = 0.008) suggests a strong internal connection. Meanwhile, the negative covariance between the Water factor and Age (-0.2235, p < .001) shows an inverse relationship, underscoring the complex dynamics involved. The model fit indices raise important concerns about the adequacy of the CFA model. The RMSEA value of 0.594 exceeds the acceptable threshold of 0.08, showing a poor fit. Although this prompts questions about the model’s specification, the Akaike Information Criterion (AIC) (6527) and the Bayesian Information Criterion (BIC) (6636) serve as useful metrics for comparing alternative models and guiding future improvements. In conclusion, the CFA results highlight the complexity of the relationships among the examined constructs. While indicators represent their respective latent factors, for the Climate Change and Age constructs, the Water factor is challenged by negative loadings on indicators. The overall model fit shows that further refinements are necessary to capture the data better.

#### Structural Equation Modelling (SEM)

The Structural Equation Modelling (SEM) analysis employed Diagonally Weighted Least Squares (DWLS) estimation, yielding valuable insights into the relationships among constructs related to child health. Despite the complexity of these relationships, the model shows a strong fit. Model fit and overall tests yielded a Chi-square value of 25.9 with 24 degrees of freedom (p = 0.358), showing no significant difference from a perfect fit, which is a positive outcome. In contrast, the Baseline Model showed a high Chi-square value of 995.9 (p < 0.001), suggesting that the User Model significantly outperformed a simple independence model. This conclusion is supported by fit indices, including a Comparative Fit Index (CFI) of 0.998 and a Tucker-Lewis Index (TLI) of 0.997, both showing excellent model fit. The Standardized Root Mean Square Residual (SRMR) was 0.057, well below the acceptable threshold of 0.08, further confirming the model’s adequacy.

The measurement model estimates revealed significant pathways between the latent constructs and their observed variables. For the Age construct, the variables sanitation needs (β = 1.192, p < 0.001) and age-specific factors (β = 0.804, p < 0.001) significantly influenced it. This suggests that increases in sanitation needs and age-specific factors are associated with higher values of the Age construct, emphasizing their importance in understanding child health dynamics. Regarding the climate change construct, both waterborne issues (β = 1.000) and heat stress (β = 1.273, p < 0.001) were significant, highlighting the role of climate-related factors in child health outcomes. The Sanitation Facilities (SF) construct also showed strong relationships with Access Sanitation (β = 1.000), Excreta facilities (β = 1.208, p < 0.001), Accessibility (β = 1.231, p < 0.001), and Excreta type (β = 0.938, p < 0.001). These results show that better access to sanitation and facilities contributes to improved health outcomes for children. The variances of the latent variables were significant, showing considerable variability across the constructs. For example, the variance for Age was estimated at 0.342 (p < 0.001), suggesting that age-related factors greatly influence the model. Notably, the covariance between Age and Climate (-0.211, p < 0.001) shows an inverse relationship; as age increases, the effect of climate factors on health outcomes decreases. This finding implies that older children have different vulnerabilities to or resilience against climate factors than younger children. The positive covariance between Age and SF (0.161, p < 0.001) suggests that as children age, the impact of sanitation facilities on child health increases, showing a potential interaction in which older children benefit more from improved sanitation practices.

In summary, the SEM analysis shows that age, climate, and access to sanitation facilities are key factors influencing child health outcomes. The model shows a strong fit, with significant relationships among the constructs. These findings underscore the importance of addressing sanitation needs and climate-related factors in health interventions to improve child health. Future research will examine additional variables or refine the model to better understand these complex interactions.

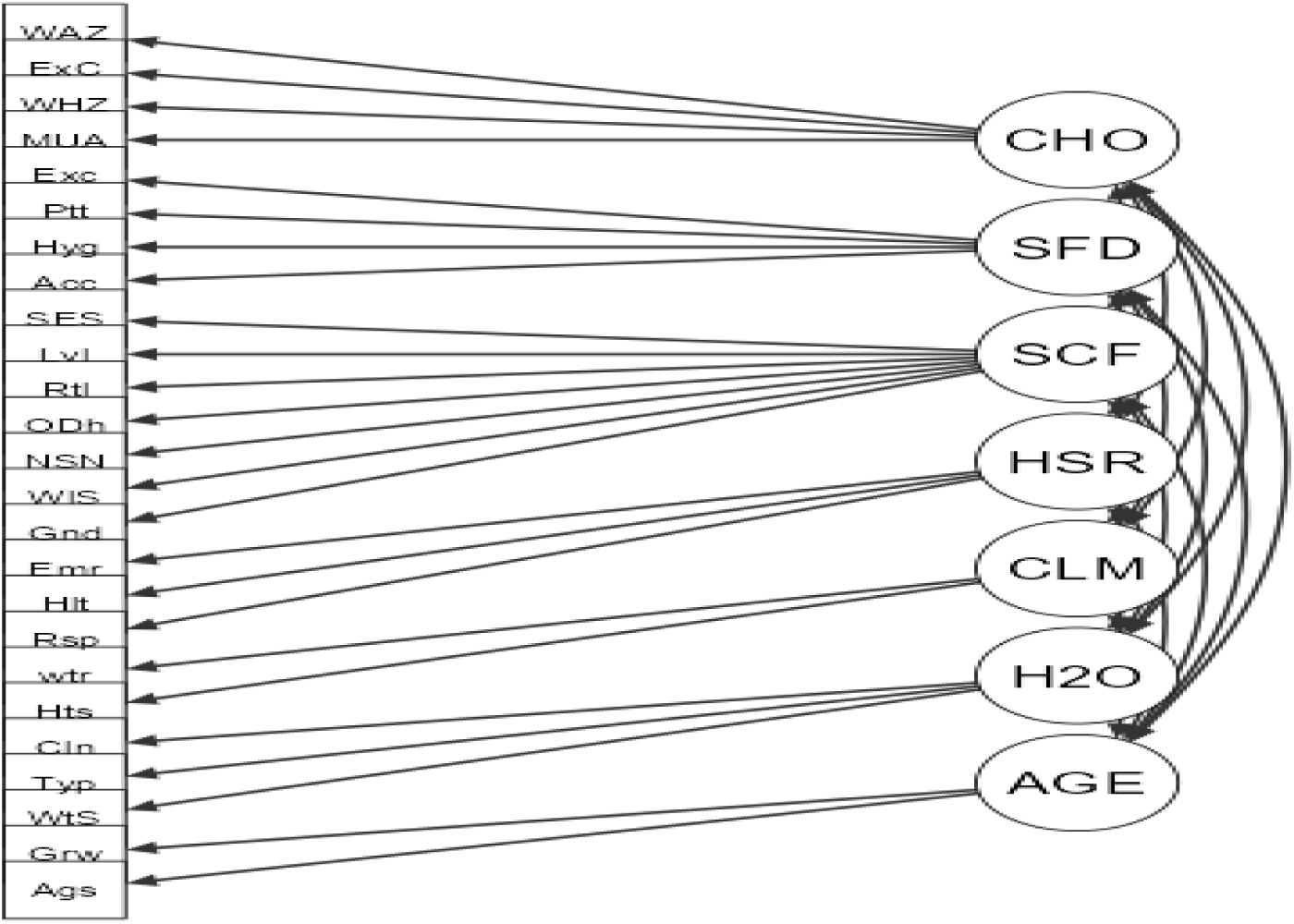
Confirmatory Factor Analysis-. Path Diagram

**Figure 2:**
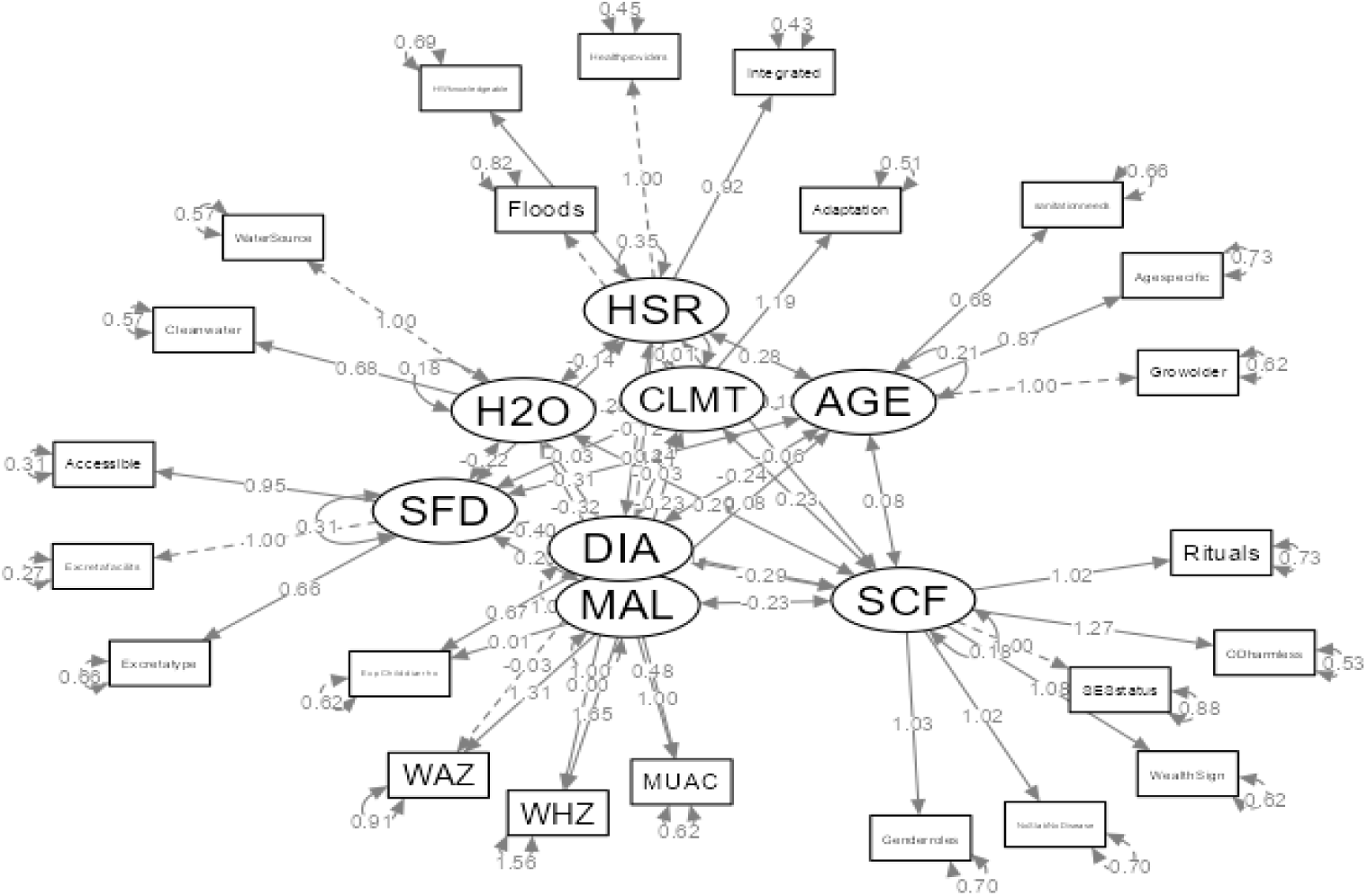
Confirmatory Factor Analysis (CFA) Findings. Key: SFD= sanitation facility & diaper; SCF=sociocultural factors; HSR=Health system response; CLMT=Climate, H20=water, DIA=Diarrhea Prevalence (DP), MAL=Malnutrition Structural Equation Modelling (SEM*)

### Regression Moderation Findings

The reliability analysis demonstrated strong internal consistency among the measured constructs, as shown by Cronbach’s alpha values: sociocultural factors (α = 0.94), sanitation facilities (α = 0.75), health facility response (α = 0.90), and child health outcomes (α = 0.76). Furthermore, the analysis verified that multicollinearity was not an issue, with tolerance values ranging from 0.647 to 1.00 and variance inflation factor (VIF) values between 1.00 and 1.735.

Hierarchical regression results, as shown in Table 4, indicate that including sociocultural factors, sanitation facilities, and health system responses explained 32% of the variance in childhood diarrhea (R² = 0.10 in Model 1, 0.13 in Model 2, and 0.14 in Model 3).

**Table 4:**
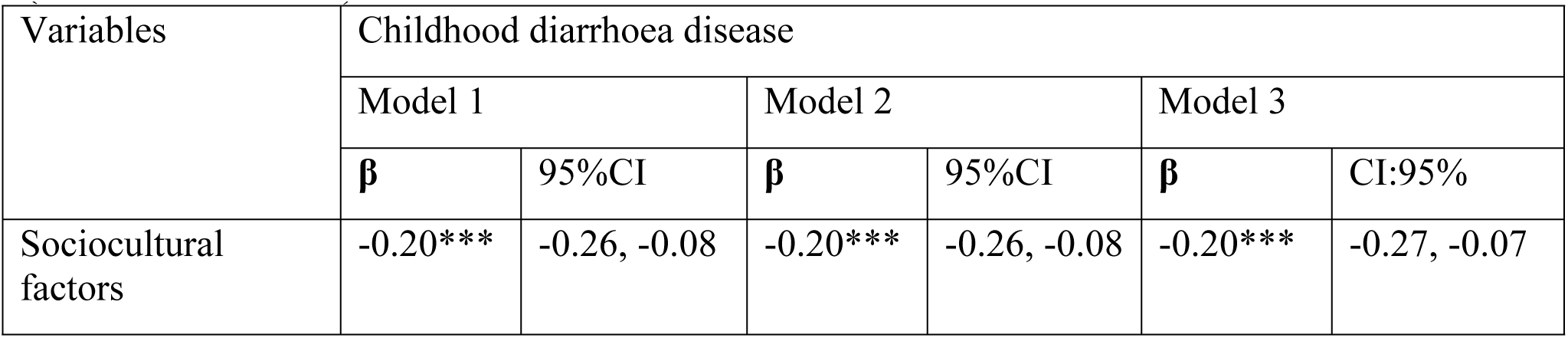
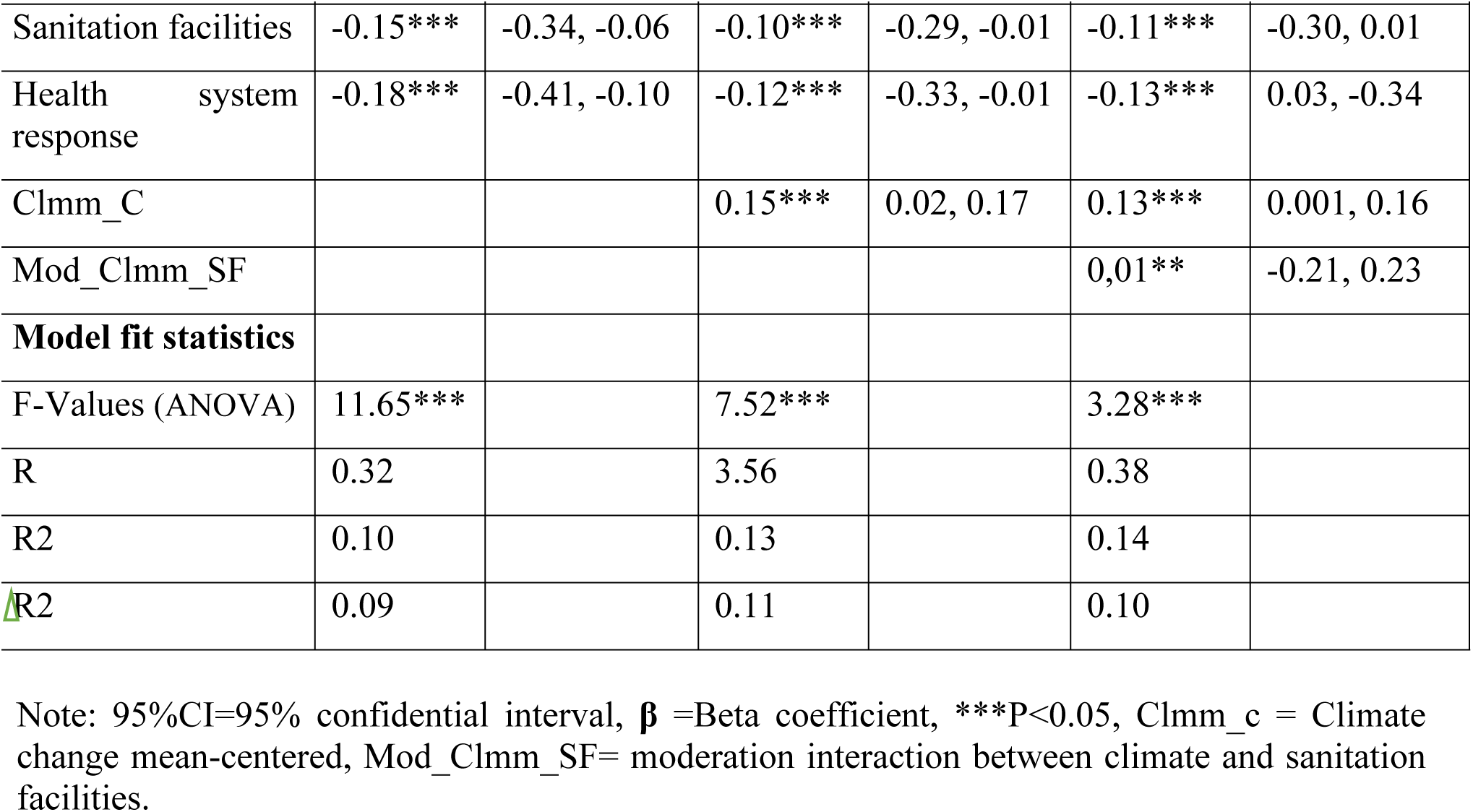
Complex Moderation Analysis of Sanitation Practices and Child Health Outcomes (Diarrheal Diseases)

From Table 4, starting with Model 1, the analysis examined the effects of sociocultural factors, sanitation facilities, and health system responses on childhood diarrhoea. The beta coefficient for sociocultural factors was -0.20 (95% CI: -0.26 to -0.08, p < 0.001), showing a significant negative relationship. This means that improvements in sociocultural conditions are associated with a decrease in diarrhoea cases. Similarly, the sanitation facilities variable showed a significant adverse effect (β = -0.15, 95% CI: -0.34 to -0.06, p < 0.001), showing that better sanitation is associated with lower diarrhoea rates. The health system response showed a strong negative association (β = -0.18, 95% CI: -0.41 to -0.10, p < 0.001), showing that a strong health system is associated with better health outcomes for children. The model fit statistics included an R² of 0.10 and an adjusted R² of 0.09, with an F-value of 11.65 (p < 0.001), confirming the model’s predictive significance.

In Model 2, including climate change factors (Clmm_C) showed a significant positive association (β = 0.15, 95% CI: 0.02 to 0.17, p < 0.001) with childhood diarrhoea outcomes, suggesting that adverse climate conditions increase diarrhoea rates, especially when sanitation practices are poor. The model’s fit statistics showed an R² of 0.13 and an adjusted R² of 0.11, with an F-value of 7.52 (p < 0.001), confirming the model’s significance. Significantly, neither age nor water access alone moderates the relationship between these factors and childhood diarrhoea outcomes.

Model 3 examines how climate change influences sanitation facilities (Mod_Clmm_SF), with a beta coefficient of 0.01 (95% CI: -0.21 to 0.23; p < 0.05). Although this interaction is statistically significant, the confidence interval shows uncertainty about the effect size, suggesting that the relationship between sanitation facilities and childhood diarrhoea varies with climate conditions. This model also confirms that climate change significantly moderates these relationships (p < 0.05), while age and access to water do not have similar moderating effects. The model’s fit statistics include an R² of 0.14 and an adjusted R² of 0.10, with an F-value of 3.28 (p < 0.001), showing strong predictive capability. High Cronbach’s alpha values for the variables in this model affirm the robustness of the results across different specifications.

The regression-moderation findings reveal the complex relationships between sociocultural factors, sanitation facilities, and health system responses in childhood diarrhoea. The significant impact of these predictors, combined with the moderating role of climate change, underscores the need for comprehensive public health strategies that address the multiple factors influencing child health outcomes.

As shown in Table 4 (Overview of Models), the analysis comprises three models that assess how various factors influence childhood diarrhoea, focusing on the moderating roles of climate change (Clmm_C) and sanitation facilities (Mod_Clmm_SF).

Table 5 under Model 1 analyses how sociocultural factors, sanitation facilities, and health system responses impact childhood malnutrition. The beta coefficient for sociocultural factors is - 0.43 (95% CI: -0.63 to -0.39, p < 0.001), showing a significant negative relationship. This suggests that improvements in sociocultural conditions are strongly associated with reduced childhood malnutrition. The sanitation facilities variable has a beta of 0.01 (95% CI: -0.16 to 0.22), suggesting no significant effect on malnutrition outcomes. The health system response shows a significant adverse effect (β = -0.13, 95% CI: -0.46 to -0.06, p < 0.001), showing that strong health system engagement correlates with lower child malnutrition. The model fit statistics report an F-value of 28.917 (p < 0.001), with an R² of 0.21 and an adjusted R² of 0.22, showing the model explains 21% of the variance in childhood malnutrition. The Cronbach’s alpha for the variables shows high reliability, though the exact value is not specified.

**Table 5:** Moderation between sanitation practices and childhood malnutrition.

| Variables | Childhood malnutrition using MUAC |  |  |  |  |  |
| --- | --- | --- | --- | --- | --- | --- |
|  | Model 1 |  | Model 2 |  | Model 3 |  |
| | $\beta$ | CI:95% | $\beta$ | CI:95% | $\beta$ | CI:95% |
| Sociocultural factors (SC) | -0.43*** | -0.63, -0.39 | -0.43*** | -0.64, -0.40 | -0.39*** | -0.60, -0.34 |
| Sanitation facilities (SF) | 0.01 | -0.16, 0.22 | 0.01 | -0.18, 0.21 | 0.04 | -0.13, 0.28 |
| Health system response (HF) | -0.13*** | -0.46, -0.06 | -0.14*** | -0.48, -0.06 | -0.14*** | -0.49, -0.60 |
| Agem_C |  |  | 0.28** | -0.10, 0.17 | -0.02 | -0.17, 0.12 |
| Mod_Age_SC interaction |  |  |  |  | 0.16*** | 0.09, 0.59 |
| Mod_Age_SF interaction |  |  |  |  | 0.10** | -0.89, 0.78 |
| Model fit statistics |  |  |  |  |  |  |
| F-Values (ANOVA) | 28.917*** |  | 14.438*** |  | 6.588*** |  |
| R | 0.47 |  | 0.47 |  | 0.50 |  |
| R<sup>2</sup> | 0.21 |  | 0.20 |  | 0.21 |  |
| ΔR<sup>2</sup> | 0.22 | | 0.00 | | 0.03 | |

Note: $\beta$ represents the Beta coefficient. \*\*\* shows statistical significance at $P < 0.05$ . Agem\_C denotes the moderator age, mean-centered; Mod\_Age\_SC signifies the interaction between moderator age and sociocultural factors; and Mod\_Age\_SF shows the interaction between moderator age and sanitation facilities.

In Model 2, including the age moderator (Agem_C) yields a beta coefficient of 0.28 (95% CI: -0.10 to 0.17, p < 0.01), showing a significant positive association between age and childhood malnutrition, suggesting that older children experience different malnutrition outcomes. Sociocultural factors remain significant (β = -0.43, 95% CI: -0.64 to -0.40, p < 0.001), and the health system response continues to have a significant adverse effect (β = -0.14, 95% CI: -0.48 to -0.06, p < 0.001). The sanitation facilities variable has a beta coefficient of 0.01 (95% CI: -0.18 to 0.21), showing no change in its effect. The model fit statistics show an F-value of 14.438 (p < 0.001), an R² of 0.20, and an adjusted R² of 0.00, showing that including the age moderator does not significantly enhance the model’s explanatory power. The reliability of the constructs remains high.

Model 3 examines the interaction effects in greater detail. The moderation interaction between age and sociocultural factors (Mod_Age_SC) shows a beta coefficient of 0.16 (95% CI: 0.09 to 0.59, p < 0.001), showing that age moderates the relationship between sociocultural factors and malnutrition and offers insights into this relationship. The moderation interaction between age and sanitation facilities (Mod_Age_SF) has a beta of 0.10 (95% CI: -0.89 to 0.78), which is not statistically significant. Sociocultural factors continue to impact malnutrition significantly negatively (β = -0.39, 95% CI: -0.60 to -0.34, p < 0.001), whereas the health system response remains significant (β = -0.14, 95% CI: -0.49 to -0.60, p < 0.001). The model fit statistics display an F-value of 6.588 (p < 0.001), an R² of 0.21, and an adjusted R² of 0.03, showing a modest improvement in model fit. The high reliability of the constructs underscores the robustness of the findings.

In summary, the findings highlight the importance of sociocultural factors and health system responses in addressing childhood malnutrition, whereas sanitation facilities have little to no impact. The moderating effect of age on the relationship between sociocultural factors and malnutrition suggests that age-specific interventions are necessary to reduce malnutrition rates.

## IV DISCUSSION

The findings highlight the complex relationships between socio-economic status, sanitation practices, and child health outcomes in Gulu District, Uganda. Consistent with existing literature (UNICEF, 2019; WHO, 2018), the results show that households with low socio-economic status are more likely to have poor sanitation practices, where 24.9% lacked access to sanitation facilities and 20.5% had handwashing facilities with soap. This is a big concern because these households are also vulnerable to climate-related shocks such as floods, which worsen sanitation issues. The prevalence of unimproved anal cleansing (68.1%) and open disposal of diapers (64.1%) highlights the need for targeted interventions to improve sanitation infrastructure and promote hygienic behaviours. The decline in sanitation practices among older children is also alarming, suggesting increased risk of sanitation-related health issues.

Diarrhoea poses a significant health risk linked to poor sanitation, in line with global estimates (WHO, 2019). The impact of climate on sanitation practices and child health outcomes is also noteworthy: Half of respondents (48.3%) reported collapsed latrines because of floods, and 63.4% cited climate as a significant factor in waterborne diseases. These results highlight the need for climate-resilient sanitation infrastructure and adaptive strategies to reduce the effects of climate change. The limited financial capacity of households to build latrines or cover medical expenses remains a critical issue, emphasizing the need for targeted financial support and social protection mechanisms. Consistent with the Sustainable Development Goals (SDGs), our findings stress the importance of ensuring universal access to affordable, sustainable sanitation services (SDG 6) and enhancing climate resilience (SDG 13).

To address the urgent sanitation issues in Gulu District, Uganda, targeted interventions, social protection mechanisms, climate-smart sanitation planning, and education and awareness campaigns are essential. This is accomplished by developing sanitation solutions tailored to specific contexts and resilient to climate change, creating financial support programs that integrate climate change considerations into sanitation planning, and starting hygiene education and awareness campaigns. The initiatives make policymakers and practitioners foster health environments, mitigate health risks, and contribute to achieving the Sustainable Development Goals (SDGs).

### Moderation Analysis between Sanitation Practices and Diarrhoea Outcomes

The moderation analysis findings provide valuable insights into the complex relationships among sociocultural factors, sanitation facilities, health system responses, and climate change in their effect on childhood diarrhoea outcomes. Abductive reasoning shows that these relationships are complex and shaped by multiple factors (Biran et al. 2022). This perspective is supported by the existing literature, which underscores the importance of considering multiple factors in understanding health outcomes (Biran et al., 2022).

In Model 1, the analysis shows a significant negative link between sociocultural factors (β = -0.20, p < 0.001) and childhood diarrhoea. This result aligns with existing research emphasizing the key role of sociocultural factors, such as community norms, education, and awareness, in improving health outcomes. For example, studies show that better education and community involvement improve hygiene practices, which are essential for reducing diarrhoea among children (Contreras et al., 2022). The strength of this finding underscores the need to incorporate sociocultural perspectives into public health strategies, aligning with the Social-Ecological Model (SEM), which promotes multilevel approaches to understanding health behaviours (Golden et al., 2015).

The analysis further reveals that sanitation facilities have a significant impact on childhood diarrhoea outcomes, with a beta coefficient of -0.15 (p < 0.001). This finding reinforces the connection between access to adequate sanitation and health outcomes. Previous studies have consistently shown that improved sanitation facilities are associated with lower rates of diarrheal disease (WHO, 2022). The SEM highlights this by emphasising the interplay between environmental conditions and health, underscoring the importance of systemic enhancements to sanitation infrastructure to improve child health.

Health system responses were significantly negatively associated with childhood diarrhoea (β = -0.18, p < 0.001). This result shows that effective health system interventions, such as timely health education and accessible healthcare services, are essential for reducing the incidence of diarrhoea. This finding aligns with previous research emphasizing the importance of robust health systems in addressing public health challenges among vulnerable groups (Pickbourn et al., 2022).

In Model 2, including climate change factors (Clmm_C) yields a positive coefficient (β = 0.15, p < 0.001), showing that adverse climate conditions exacerbate childhood diarrhoea in the presence of poor sanitation. This finding is important as it aligns with the increasing recognition of climate change as a public health issue. Studies suggest that climate-related factors, such as extreme weather events and temperature changes, significantly affect water quality and sanitation, increasing the risk of diarrheal diseases (Shrestha et al., 2020). The SEM framework supports this view by illustrating how environmental changes influence health outcomes, highlighting the need for flexible public health strategies in response to climate variability.

Model 3 examines the moderation effect of climate change on sanitation facilities (Mod_Clmm_SF), showing a beta coefficient of 0.01 (p < 0.05). Although this interaction is significant, the confidence interval shows uncertainty about the magnitude of the effect. This shows that the link between sanitation facilities and childhood diarrhoea depends on climate conditions. This complexity is consistent with the Diffusion of Innovations Theory, which states that the success of health innovations, such as improved sanitation, is affected by external factors, including environmental conditions and community readiness to adopt new practices (Rogers, 2003).

Overall, the findings from this moderation analysis highlight the complex interplay between sociocultural factors, sanitation practices, and health system responses in shaping childhood diarrhoea outcomes. The significant effects of these predictors, along with the moderating role of climate change, underscore the need for integrated public health strategies that address multiple determinants of child health. Such strategies will be guided by theoretical frameworks, such as SEM and the diffusion of innovations theory, which promote holistic approaches that account for the social and environmental context when addressing health challenges.

The analysis emphasizes that addressing childhood diarrhoea requires a multifaceted approach that includes improving sociocultural conditions, enhancing sanitation facilities, and strengthening health system responses, while accounting for the impacts of climate change. This comprehensive understanding is crucial for designing effective public health interventions to ease the burden of childhood diarrhoea and enhance overall child health outcomes.

### Moderation Analysis between Sanitation Practices and Child Malnutrition Outcomes

The results of the moderation analysis on childhood malnutrition are significant. They offer valuable insights into how sociocultural factors, sanitation facilities, health system responses, and age interact. This analysis, which examines how these variables contribute to childhood malnutrition, addresses an urgent public health issue that warrants global attention.

In Model 1, the analysis shows a strong negative relationship between sociocultural factors and childhood malnutrition (β = -0.43, p < 0.001). This suggests that improvements in sociocultural conditions, such as community involvement, education, and awareness, are linked to lower rates of child malnutrition. This finding aligns with existing research showing that sociocultural contexts influence dietary practices and health behaviours. For example, Shrestha et al. (2022) show that sociocultural dynamics, including food preferences and family feeding practices, are vital for addressing malnutrition in West and Central Africa. Ummah (2019) and Bauza et al.(2019) also highlight that social and economic factors affect household food distribution, which impacts children’s nutritional intake. These studies reinforce the idea that strengthening sociocultural factors leads to better nutritional outcomes for children.

Interestingly, the sanitation facilities variable in Model 1 shows no significant effect on malnutrition outcomes (β = 0.01, p > 0.05). This finding differs from literature that emphasizes the critical role of sanitation in child health. However, it suggests that sanitation’s direct impact on malnutrition compared to sociocultural factors. This aligns with Social Learning Theory (SLT), which states that behaviours are learned through observation and imitation. In this context, if sociocultural norms do not prioritize proper sanitation, improving sanitation facilities will not reduce malnutrition rates.

The health system response also shows a significant adverse effect (β = -0.13, p < 0.001). This highlights the crucial role of health system engagement in fighting childhood malnutrition, consistent with the literature, which supports strong health systems that provide effective nutritional interventions. For example, UNICEF (2023) examines how cash transfer programs and health interventions help reduce malnutrition among vulnerable groups, emphasizing that resilient health systems must address malnutrition.

In Model 2, adding age as a moderator reveals a significant positive link between age and childhood malnutrition (β = 0.28, p < 0.01). This suggests that older children experience different outcomes from malnutrition due to changing dietary needs and exposure to diverse sociocultural environments as they grow. The persistent significance of sociocultural factors (β = -0.43) and the health system response (β = -0.14) in this model underscores their continued relevance across age groups. This suggests the potential for targeted interventions that incorporate age-related factors into nutritional programs, improving outcomes.

Model 3 further examines interaction effects between age and sociocultural factors (Mod_Age_SC), with a beta coefficient of 0.16 (p < 0.001). This shows that the influence of sociocultural factors on malnutrition is moderated by age, suggesting that strategies to improve sociocultural conditions need to be tailored for different age groups. In contrast, the interaction between age and sanitation facilities (Mod_Age_SF) is not significant, suggesting that sanitation alone may not address age-related disparities in malnutrition outcomes.

In summary, the analysis emphasizes the importance of comprehensive, tailored public health strategies that account for the complex nature of childhood malnutrition. The findings from the moderation analysis, guided by theoretical frameworks such as the Social-Ecological Model (SEM) and the Diffusion of Innovations Theory, provide a comprehensive understanding of the factors that affect childhood malnutrition, showing the rigor of the research.

Limitations: The moderation analysis in this study is subject to limitations. First, the complex interactions among sociocultural factors, sanitation practices, and health system responses do not encompass determinants, leaving gaps in understanding the full range of determinants of childhood health outcomes. The considerable uncertainty in interaction effects between climate change and sanitation facilities suggests that unmeasured factors also be important, limiting the extent to which the findings be generalized. The lack of significant impacts from sanitation facilities on childhood malnutrition suggests a complex relationship, showing that sanitation’s influence on nutrition may not be as straightforward as once thought.

The context-specific nature of the results highlights sociocultural dynamics varying across populations and regions, which limits the extent to which the findings always applicable. Although age was recognized as a moderator in the analysis, this adds complexity to understanding how age-related factors influence access to resources and health behaviours, underscoring the need for detailed, age-targeted interventions. Reliance on existing literature and data introduces inherent biases that impact the validity of the results. The analysis reflects short-term associations rather than long-term trends, suggesting that longitudinal data provide a complete understanding of how these factors affect childhood health outcomes. Recognizing these long-term patterns is vital to developing effective, sustainable strategies to reduce childhood malnutrition rates. Addressing these limitations is crucial for improving public health approaches and shaping future research efforts.

Addressing childhood malnutrition requires a nuanced understanding of how sociocultural factors, health system responses, and age interact. Targeted interventions that account for these variables are crucial for reducing malnutrition rates and improving child health outcomes. The results from the moderation analysis on childhood diarrhoea and malnutrition highlight the need for specific policy changes, future research directions, and guidance for decision-makers.

### Final interpretation of the moderation analysis

The moderation analysis uncovers complex links between sociocultural factors, sanitation facilities, health system responses, and climate change that affect childhood diarrhoea and malnutrition outcomes. Key findings include a notable decrease in childhood diarrhoea related to sociocultural factors (β = -0.20, p < 0.001) and access to sanitation facilities (β = -0.15, p < 0.001). Climate change worsens childhood diarrhoea, especially when sanitation practices are poor (β = 0.15, p < 0.001). This shows that climate change influences childhood health outcomes by affecting factors like sanitation. The effect of sanitation facilities on childhood diarrhoea depends on climate conditions.

Regarding childhood malnutrition outcomes, sociocultural factors significantly reduce malnutrition rates (β = -0.43, p < 0.001), and health system responses have a notable effect on malnutrition outcomes (β = -0.13, p < 0.001). Age influences the relationship between sociocultural factors and childhood malnutrition, underscoring the need for tailored interventions across age groups. However, sanitation facilities do not appear to have a significant direct impact on childhood malnutrition outcomes.

The findings of this study have theoretical implications. The Social-Ecological Model (SEM) and Diffusion of Innovations Theory aligns to these findings, emphasizing the importance of considering multiple factors and contextual influences (Golden et al., 2015; Rogers, 2003). The analysis highlights the need for comprehensive public health strategies that address sociocultural, environmental, and health system factors.

The study’s findings also have policy implications. Targeted interventions will focus on improving sociocultural conditions, enhancing sanitation facilities, and strengthening health system responses to tackle these challenges. Climate change mitigation strategies will be incorporated into public health policies to address the worsening effects of childhood diarrhoea. Age-specific interventions are essential for tackling childhood malnutrition.

Future research directions include investigating the impact of climate change on childhood health outcomes across contexts, examining the role of sanitation facilities in addressing childhood malnutrition, and developing and evaluating age-specific interventions to prevent and treat it.

### Final Summary

This study examined the interactions among sociocultural factors, sanitation facilities, health system responses, and climate change in their effects on childhood diarrhoea and malnutrition outcomes. The moderation analysis revealed complex relationships among these variables, underscoring the need to account for multiple factors and contextual influences. The findings show that sociocultural factors, sanitation facilities, and health system responses are crucial in reducing childhood diarrhoea and malnutrition. Climate change exacerbates childhood diarrhoea, whereas age moderates the relationship between sociocultural factors and childhood malnutrition.

This study provides valuable insights into the complex relationships among sociocultural factors, sanitation facilities, health system responses, and climate change in shaping childhood diarrhoea and malnutrition outcomes. The findings highlight the need for integrated public health strategies that address multiple aspects of child health. Policymakers, practitioners, and researchers will recognize the intricate interactions between these variables when developing interventions to combat childhood diarrhoea and malnutrition. Tackling these urgent public health issues requires a comprehensive and nuanced approach that considers the social, environmental, and healthcare contexts in which children live.

### Contribution to the body of knowledge

This study makes a significant contribution to the existing knowledge base by improving our understanding of the sociocultural factors that impact childhood health outcomes. The research provides detailed insights into the complex relationship between sociocultural factors, sanitation practices, and health system responses in shaping childhood health. Notably, the findings highlight the vital role of sociocultural conditions in decreasing the rates of childhood diarrhoea and malnutrition.

The study’s results also clarify how climate change and age influence the relationships between sanitation, sociocultural factors, and health outcomes. This understanding deepens the understanding of how these variables are connected. By highlighting the moderating effects of climate change and age, this study supports the development of targeted interventions that address the specific challenges posed by environmental shifts across different age groups.

The research findings highlight the importance of comprehensive strategies that include sociocultural factors, robust health system efforts, and sanitation improvements. This knowledge helps develop more effective, tailored interventions to reduce childhood diarrhoea and malnutrition. By promoting a multifaceted approach, this study underscores the need to address the complex interplay among sociocultural influences, sanitation practices, and health system responses.

The study’s use of the Social-Ecological Model (Golden et al., 2015) and Diffusion of Innovations Theory (Rogers, 2003) enhances understanding of the complex connections among individual, relational, community, and societal factors affecting health outcomes. By expanding these frameworks, this research offers a detailed view of how sociocultural factors, sanitation practices, and health system responses are interconnected.

The research findings offer practical insights for policymakers, public health practitioners, and community leaders to design and implement effective interventions for addressing the complex challenges of childhood diarrhoea and malnutrition. In summary, this study advances our knowledge by deepening our understanding of the intricate relationship between sociocultural factors, sanitation practices, and health system responses. The findings guide the development of comprehensive interventions, expand theoretical frameworks, and improve practical applications, leading to better health outcomes for children worldwide.

This study adds to the existing knowledge by deepening our understanding of how sociocultural factors, sanitation practices, and health system responses interact. The results guide the creation of comprehensive interventions, expand theoretical frameworks, and improve practical applications, leading to better health outcomes for children worldwide.

## V CONCLUSION AND RECOMMENDATION

This study’s moderation analysis of childhood diarrhoea and malnutrition emphasizes the complex interactions among sociocultural factors, sanitation practices, and health system responses, as well as the moderating effects of climate change and age. The findings confirm that improved sociocultural conditions are linked to lower rates of childhood diarrhoea, underscoring the important role of community engagement and education in health outcomes.

Access to proper sanitation facilities is vital, as effective sanitation practices are associated with lower rates of diarrheal diseases (WHO, 2025). Health system responses are crucial, as strong healthcare interventions reduce the impact of diarrhoea in children.

The analysis of childhood malnutrition shows that sociocultural factors are crucial, with stronger community dynamics and awareness associated with lower malnutrition rates. The absence of a significant direct effect of sanitation facilities on malnutrition outcomes suggests sociocultural norms overshadow the benefits of sanitation improvements, consistent with Social Learning Theory (SLT) (Bandura, 2001). The findings emphasize the importance of health system engagement in addressing malnutrition, as targeted interventions improve nutritional outcomes. The analysis confirms that addressing childhood health issues requires comprehensive, integrated public health strategies.

These strategies will focus on improving sociocultural conditions, sanitation facilities, and health system responses, while accounting for the effects of climate change and population aging. The Social-Ecological Model (SEM) provides a comprehensive framework for understanding the multiple levels that influence health outcomes (Golden et al., 2015). In contrast, the Diffusion of Innovations Model (DIM) emphasizes the importance of effective communication and promotion of health innovations within communities (Rogers, 2003).

The study recommends that policymakers develop community-centered health strategies that account for sociocultural and climatic factors and that improve sanitation. Community leaders promote health education and water management programs to boost overall well-being. Health practitioners will tailor interventions to different age groups, especially for children under 24 months. Households will prioritize hygiene and work with local leaders and health workers. Researchers are encouraged to perform longitudinal studies on sociocultural influences and age-specific interventions.

## Data Availability

Data will be available on request

## Abbreviations

CI: Confidential Interval;
EFA: Exploratory Factor Analysis;
CFA: Confirmatory Factor Analysis;
SEM: Structural Equation Modelling;
KI: Key Informants;
FGD: Focus Group Discussion;
COR: Crude Odds Ratio;
AOR: Adjusted Odds Ratio;
QDA: Qualitative Data Analysis;
CLTS: Community-Led Total Sanitation;
SFD: Sanitation Facilities and Diapers;
SCF: Sociocultural Factors;
HSF: Health System Factors;
ODF: Open Defecation Free.

## Declarations

### Ethical approval and consent from participants

The research study was conducted with the highest regard for ethical principles. The Helsinki Declaration minimized harm and maximized benefits for participants. Before starting the study, ethical approval was secured from the Lacor Hospital Institutional Review Board (LHIRB) (reference number LACOR-2014-347) and the Uganda National Council for Science and Technology (UNCST) (reference number HS4991ES). The LHIRB reviewed and approved the research protocol, ensuring ethical standards were maintained throughout the study.

Before participating in the study, participants received a detailed information sheet explaining the study’s purpose, risks, and benefits. They were also informed of their right to withdraw without penalty. Written consent was obtained from participants, and their confidentiality and anonymity were maintained throughout the study.

Compliance with the Helsinki Declaration throughout the study ensured that the principles outlined were maintained. Participants’ autonomy and dignity were honoured, and their rights and well-being were safeguarded. The researchers recognized their responsibilities to the participants, the institution, and the broader community and took steps to ensure that the study was conducted with integrity and transparency.

### Consent for publication

Not applicable

### Availability of data and materials

Data and materials used in this study are available from the editor upon request, promoting transparency and facilitating further research. The corresponding author is also available to provide any additional information or clarification.

### Authors Contributions

YI, a key contributor, played an important role in the study’s conception and design, created the questionnaire, led data collection and analysis, and drafted the initial manuscript. NDN and AB, as co-authors, provided their expertise by reviewing and approving the manuscript for submission, enhancing the study’s credibility.

## Acknowledgment

I extend my heartfelt appreciation to the research assistants, who were essential to the collection of both quantitative and qualitative data. Their dedication and hard work have been invaluable to this study. I also acknowledge the district leadership for their unwavering support, as well as Mshilla Maghanga and the reviewers of this work.

## Author Details

Directorate of Postgraduate Studies and Research, Nkumba University, Uganda, P.O. Box 237, Entebbe, Uganda.

